# Inactive disease in lupus patients is linked to autoantibodies to type-I interferons that normalize blood IFNα and B cell subsets

**DOI:** 10.1101/2021.04.07.21255049

**Authors:** Madhvi Menon, Hannah F. Bradford, Liis Haljasmagi, Martti Vanker, Pärt Peterson, Chris Wincup, Rym Abida, Raquel Fernandez Gonzalez, Vincent Bondet, Darragh Duffy, David A. Isenberg, Kai Kisand, Claudia Mauri

**Author notes:** joint first authors (Madhvi Menon, Hannah F. Bradford, Liis Haljasmagi). joint senior authors (Kai Kisand, Claudia Mauri).

## Abstract

Systemic Lupus Erythematosus (SLE) is characterized by a prominent increase in expression of type-I interferon (IFN)-regulated genes in 50-75% of patients. Here we investigate the presence of autoantibodies (auto-Abs) against type I IFN in SLE patients and their possible role in controlling disease severity. We report that out of 491 SLE patients, 66 had detectable anti-IFNα-auto-Abs. The presence of neutralizing anti-IFNα-auto-Abs correlates with lower levels of circulating IFNα protein, inhibition of IFN down-stream signalling molecules and gene signatures and with an inactive global disease score. Previously reported B cell frequency abnormalities, found to be involved in SLE pathogenesis, including increased levels of immature, double negative and plasmablast B cell populations were partially normalized in patients with neutralising anti-IFNα-auto-Abs compared to other patient groups. We also show that sera from SLE patients with neutralising anti-IFNα-auto-Abs biases *in vitro* B cell differentiation towards classical memory phenotype, while sera from patients without anti-IFNα-Abs drives plasmablasts differentiation. Our findings support a role for neutralising anti-IFNα-auto-Abs in controlling SLE pathogenesis and highlight their potential efficacy as novel therapy.

## Main

Systemic Lupus Erythematosus (SLE) is an autoimmune disease with a largely heterogeneous presentation of symptoms and pathology that can affect multiple organ systems. Abnormal B cell proportions including expansion of atypical memory, also known as double negative (DN) B cells, and autoantibody-secreting plasma cells have been shown to contribute to autoimmune inflammation and tissue injury^1–3^. In addition to B cell dysfunction, between 50-75% of SLE patients present an upregulation of type-I interferon (IFN-I)-stimulated genes (ISGs) that directly correlate with disease severity and serum IFNα levels^4,5^. Type I IFN is critical for balancing immunity for optimal protection against viral infection and minimizing potential collateral damage caused by a hyperactivated immune system. We and others have previously shown that a finely tuned IFNα response is required to induce the differentiation of immature B cells into plasma cells that produce antibodies against the initial trigger as well as regulatory B cells (Bregs) that restore homeostasis^6,7^. In SLE, chronic levels of IFNα production fuel autoimmunity by promoting the differentiation of monocytes to dendritic cells (DCs)^8,9^ that activate autoreactive T cells, the generation of effector and memory CD8^+^T cells^10–12^ and by promoting the differentiation of B cells into autoantibody-producing plasma cells but not Bregs^6,13^. Clinical observations report that patients treated with IFN-I for cancer and chronic infections develop a lupus-like disease and/or anti-dsDNA antibodies^14,15^, further substantiating the pathogenic role of IFNα in lupus.

Neutralizing auto-Abs to IFN-I have been previously reported to develop in patients treated with IFNα2 or IFNβ therapy^16,17^, in the majority of patients with autoimmune polyendocrinopathy syndrome type I (APS-1)^18,19^, thymoma^20^, at lower frequencies in rheumatic diseases, including in a small cohort of lupus patients^21,22^, and more recently in a subset of patients with life-threatening COVID-19^23,24^. To date, there have been no large-scale analyses of subtype-specific anti-IFN-I-auto-Abs in SLE patients or their association with disease severity, IFNα concentration, and their role in modulating B cell responses.

To evaluate whether SLE patients develop endogenous auto-Abs to cytokines, we tested sera from 474 SLE patients and 312 healthy controls for auto-Abs against various cytokines with the Luciferase Immunoprecipitation system (LIPS) assay (clinical characteristic, genders, ethnicity are reported in Table 1). The auto-Abs to cytokines were measured in groups that included an IFNα pool (IFNα1, IFNα2, IFNα8, IFNα21), IFNω, IFNγ, IFNβ1, a T helper (Th)17 pool, (IL-17A, IL-17F, IL-22), an IFNλ pool (IL-28A, IL-28B, IL-29), an interleukin (IL) pool (IL-6, IL-7, IL-10, IL-15), and a tumor necrosis factor (TNF) pool (TNF, LTA, BAFF, APRIL) (Fig.1a, Table 2). The majority of the auto-Abs to cytokines were either undetectable or were produced at a very low concentration in patients or control sera. However, we detected a significant increase in auto-Abs to IFNα (66 out of 474 patients) and IFNω (59 out 474 patients) in SLE patients compared to healthy volunteers (Fig. 1b, c). Reactivity towards IFN-I subtypes was partially overlapping as 12% (n=43) of patients have auto-Abs to both IFNα and IFNω, whereas anti-IFNα or –IFNω single positive patients comprise 4% each, moreover we show that the levels of auto-anti-IFNα positively correlate with auto-anti-IFNω Abs (Fig. 1d). Due to the well-established pathogenic role of IFNα in promoting SLE pathogenesis, we focus our attention on the cohort of patients that display anti-IFNα-auto-Abs. IFNα-auto-Abs are predominantly of the IgG1 subclass (Fig. 1e).

**Table 1.** Demographic and clinical characteristics of SLE patients with neutralizing or non-neutralizing anti-IFNα-auto-Abs, anti-IFNα-auto-Ab^-^ patients and healthy controls.

|  | Healthy (n=312) | Cross-sectional Ab<sup>neg</sup><br>(n=425) | Cross-sectional Ab<sup>non-neut</sup><br>(n=45) | Cross-sectional Ab<sup>neut</sup><br>(n=21) |
| --- | --- | --- | --- | --- |
| Age (Range) | 66.0 (31.0-87.0) | 45.5 (17-86) | 45.1 (23-73) | 47.5 (28-72) |
| Age at Diagnosis (Range) | - | 29.1 (1-75) | 29.5 (12-63) | 28.4 (8-51) |
| Sex (female:male) | (146:166) | (391:34) | (40:5) | (19:2) |
| Sex (%)<br>female:male) | (47.8:53.2) | (92.0:8.0) | (88.9:11.1) | (90.5:9.5) |
| Ethnicity (%)<br>AC/C/SA/EA/O) | (0/100/0/0/0) | (17.9/60.9/13.4/4.9/2.8) | (35.6/48.9/13.3/2.2/0.0)) | (47.6/38.1/4.8/9.5/0) |
| Treatment (%) |  |  |  |  |
| HCQ |  | 49.4 | 35.6 | 38.1 |
| Pred |  | 50.1 | 68.9 | 42.9 |
| MTX |  | 3.8 | 6.7 | 0.0 |
| MMF |  | 9.6 | 22.2 | 14.3 |
| Aza |  | 16.2 | 17.8 | 4.8 |
| Organ involvement (A/B/C/D/E) (%) |  |  |  |  |
| Gen |  | (0.0/4.2/16.1/69.8/10.0) | (2.6/0.0/38.5/59.0/0.0) | (0.0/0.0/33.3/61.9/4.8) |
| Mucocutaneous |  | (0.6/12.2/20.0/59.3/8.0) | (2.6/7.7/5.1/82.1/2.6) | (0.0/0.0/9.5/90.5/0.0) |
| CNS |  | (0.6/1.1/4.7/60.7/33.0) | (5.1/0.0/5.1/66.7/23.1) | (0.0/0.0/0.0/76.2/23.8) |
| CVS |  | (0.3/0.6/2.8/68.4/28.0) | (0.0/2.6/2.6/61.5/33.3) | (0.0/4.8/0/57.1/38.1) |
| Mu/Skl |  | (1.4/4.7/23.3/65.1/5.5) | (0.0/7.7/15.4/77.0/0.0) | (4.8/0/19.0/71.4/4.8) |
| Vasc |  | (0.0/0.9/28.4/47.4/23.3) | (0.0/0.0/6.7/30.0/63.3) | (0.0/0.0/0.0/35.3/64.7) |
| Renal |  | (0.3/10.8/16.3/59.0/13.6) | (0.0/15.4/5.1/33.3/46.2) | (0.0/9.5/4.8/42.9/42.9) |
| Haem |  | (0.3/8.6/43.2/43.5/4.4) | (2.6/2.6/59.0/35.9/0.0) | (0.0/0.0/47.4/52.6/0.0) |

Patients fulfilling the revised classification criteria for SLE were assessed for disease activity with the British Isles Lupus Assessment Group Index (BILAG). The BILAG index is a clinical measure of disease that distinguishes activity in nine different organ systems. Each organ system was given a grade A, B, C, D or E, where A was the most active and E as least active. In this study the grades were converted into numerical scores using the BILAG-2004 index, where A=12, B=8, C=1, D=0 and E=0. Following this, global BILAG scores were calculated by adding the sum of the values from all organ systems. Patients with a global score higher than 6 were considered active. The following abbreviations are used: African-Caribbean (AC), Caucasian (C), South Asian (SA), East Asian (EA), Other (O), hydroxychloroquine (HCQ), prednisolone (Pred), methotrexate (MTX), mycophenolate mofetil (MMF), azathioprine (Aza), general (Gen) central nervous system (CNS), cardiovascular (CVS), musculoskeletal (Mu/Skl), vascular (Vasc), haematological (Haem).

**Table 2.** Prevalence of anti-cytokine auto-antibodies in SLE patients and healthy controls.

| Seroactivity | Patients (%*,n*)<br>(*n*=474) | Controls (%*,n*)<br>(*n*=312) | Fisher's<br>exact test: |
| --- | --- | --- | --- |
| IFN-α pool | 14 (66) | 2.2 (7) | P < 0.0001 |
| IFN-ω | 12 (59) | 1.0 (3) | P < 0.0001 |
| IFN-λ pool | 2 (8) | 1.0 (3) | P = 0.5404 |
| IFN-β | 1 (5) | 0 (0) | P = 0.1631 |
| IFN-γ | 4 (18) | 1.9 (6) | P = 0.2025 |
| Th-17 pool | 6 (28) | 1.6 (5) | P = 0.0031 |
| IL pool | 6 (28) | 0.3 (1) | P < 0.0001 |
| TNF pool | 7 (35) | 0.3 (1) | P < 0.0001 |
| IL-1 pool | 1 (4) | 2.9 (9) | P = 0.0421 |
| None | 47 (223) | 88.8 (277) | P < 0.0001 |

474 SLE patients and 312 healthy controls tested for auto-Abs against various cytokines with the Luciferase Immunoprecipitation system (LIPS) assay. The auto-Abs to cytokines were measured in groups that included an IFNα pool (IFNα1, IFNα2, IFNα8, IFNα21), IFNω , IFNγ , IFNβ1, a T helper (Th)17 pool, (IL-17A, IL-17F, IL-22), an IFNλ pool (IL-28A, IL-28B, IL-29), an interleukin (IL) pool (IL-6, IL-7, IL-10, IL-15), and a tumor necrosis factor (TNF) pool (TNF, LTA, BAFF, APRIL).

**Fig. 1:**
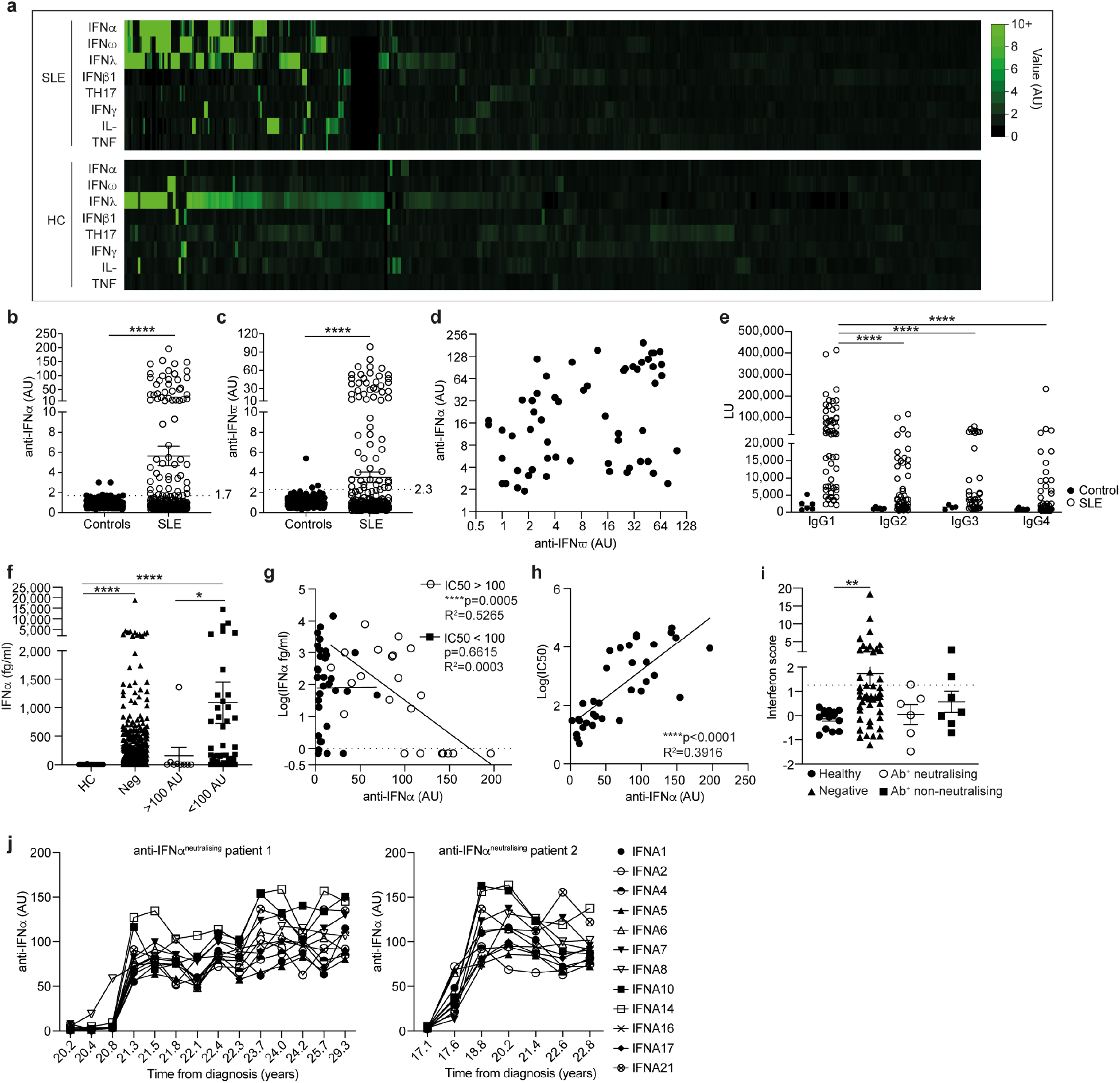
Neutralizing anti-IFNα autoantibodies in SLE patients inversely correlate with circulating IFNα. **a**, Heatmap showing levels (arbitrary units (AU)) of anti-cytokine autoantibodies (auto-Abs) (IFNα pool, IFNω, IFNλ pool, IFNβ1, Th17 pool, IFNγ, IL-pool and TNF) in 476 SLE patients and 315 healthy controls, measured by luciferase immunoprecipitation system (LIPS) assay. **b-c**, Levels of **(b)** anti-IFNα (positive cut-off 1.7 AU) and **(c)** anti-IFNω auto-antibodies (positive cut-off 2.3 AU) in sera from 491 SLE patients and 312 healthy controls. **d**, correlation between serum titres of anti-IFNα and anti-IFNω-auto-Abs in auto-Ab positive patients. \*\*\*\**P*<0.0001 by unpaired two-tailed Student’s *t* test with Welch’s correction. **e**, IgG subclasses of anti-IFNα-auto-Abs represented as luciferase units (LU) for SLE patients (n=59) and healthy controls (n=6). \*\*\*\**P*<0.0001 by non-parametric Kruskal-Wallis test with Dunn’s multiple comparison. **f**, Serum IFNα concentration of SLE patients with high (>100 AU), low (<100) or negative (<1.9 AU) titres of anti-IFNα-auto-Abs and healthy controls. \**P*<0.05, \*\*\*\**P*<0.0001 by non-parametric Kruskal-Wallis test with Dunn’s multiple comparison. **g**, Correlation between serum IFNα concentrations (measured by Simoa assay) and anti-IFNα-auto-Ab titres for SLE patients grouped according to neutralization capacity (neutralizing IC50>100, non-neutralizing IC50<100).\*\**P*=0.0024 by two-tailed nonparametric Spearman correlation. **h**, Correlation between IC50 and titre of neutralizing anti-IFNα-auto-Abs. \*\*\*\**P*<0.0001 by two-tailed nonparametric Spearman correlation. **i**, Interferon score of PBMCs isolated *ex vivo* from patients with neutralizing (n=6) or non-neutralizing (n=7) anti-IFNα-auto-Abs, anti-IFNα-auto-Ab negative SLE patients (n=47) and healthy controls (n=14). Data represented are a cumulative score of the expression of ISGs MX1, MCL1, IRF9 and STAT1 measured by RT-qPCR and relative to GAPDH. \*\**P*=0.0052 by non-parametric Kruskal-Wallis test with Dunn’s multiple comparison. **j**, Titres of neutralizing anti-IFNα-auto-Abs against IFNα subtypes longitudinally for 2 SLE patients. Error bars are shown as mean±SEM.

Next, we quantified serum IFNα levels with the ultrasensitive Simoa method^25^ and found that 93% of SLE patients had IFNα serum levels over the detection limit (0.7 fg/ml) compared to 30% of healthy controls (Supplementary Fig. 1a). Patients with high titres of anti-IFNα-auto-Abs show a significant reduction in the levels of circulating IFNα compared to anti-IFNα-auto-Ab negative and to those with low anti-IFNα-auto-Abs titres (Fig. 1f). The capacity of anti-IFNα-auto-Abs to neutralise IFNα was assessed using a reporter cell line based neutralization assay as previously described^26^. Serum levels of IFNα negatively correlated with anti-IFNα-auto-Abs titres in patients where the neutralizing capacity is IC50>100 (Fig. 1g). We observed that serum samples with high anti-IFNα-auto-Abs levels were more efficient in blocking all tested subtypes (IFNα2/5/6/8) of IFNα bioactivity *in vitro* (Fig. 1h, Supplementary Fig. 1b).

To gain mechanistic insight into the capacity of neutralizing anti-IFNα-auto-Abs to reduce downstream IFN-I signalling, we compared the IFN composite score^27^, a cumulative measure of mRNA expression of four individual ISGs: MX1, MCL1, IRF9 and STAT1 genes (see methods), in SLE patients with and without anti-IFNα-auto-Abs and healthy controls. The IFN-I score was significantly higher in anti-IFNα-auto-Ab negative patients compared to healthy controls; anti-IFNα-auto-Ab positive patients displayed an IFN-I score comparable to healthy controls (Fig. 1i, Supplementary Fig. 1c).

To measure the stability of the neutralizing anti-IFNα-auto-Abs, we selected five auto-Abs high patients from the cross-sectional group and measured the titre and neutralization capacity of these auto-Abs longitudinally over an average of 10 years from the first sample collection. All patients tested have auto-Abs against 12 subtypes of IFNα (IFNα1, IFNα2, IFNα4, IFNα5, IFNα6, IFNα7, IFNα8, IFNα10, IFNα14, IFNα16, IFNα17 and IFNα21) at high titres. Interestingly, two of the patients developed anti-IFNα-auto-Abs during follow-up analysis; one patient initially developed auto-Abs against IFNα8, and subsequently developed cross-reactivity to all the subtypes during following months (Fig. 1j and Supplementary Fig. 1d).

We next investigated the effect of neutralizing anti-IFNα-auto-Abs on disease severity. We observed that patients with at least one neutralizing antibody against an IFNα subtype displayed significantly lower disease activity (as measured by the global British Isles Lupus Assessment Group (BILAG) score) compared to patients without Abs in circulation or to those displaying Abs with negligible neutralizing capacity (Fig. 2a).

**Fig. 2:**
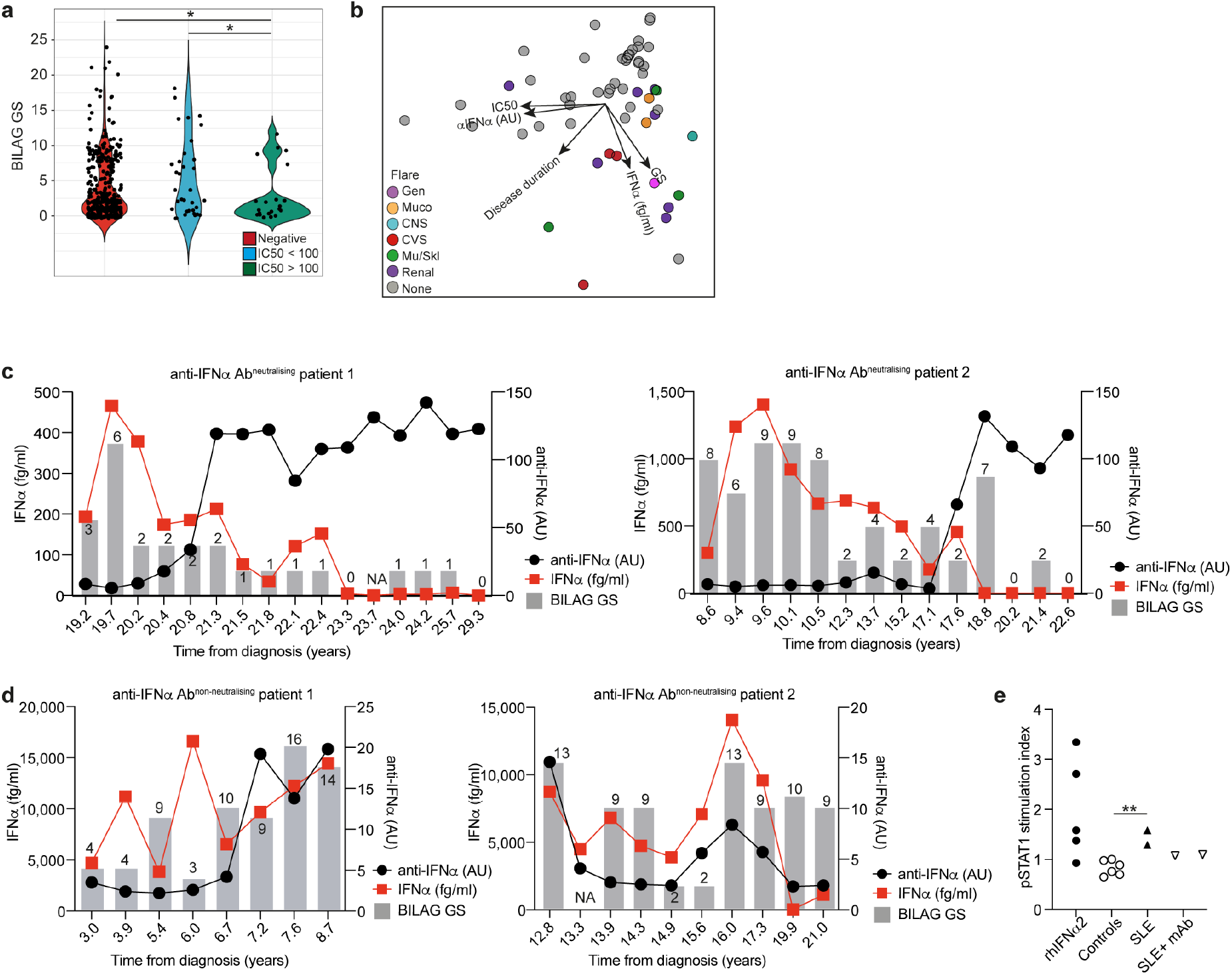
Neutralizing anti-IFNα autoantibodies are longitudinally stable, neutralize IFNα *in vivo*, and are associated with inactive disease. **a**, Violin plots show disease activity as assessed by British Isles Lupus Assessment Group (BILAG) global score (GS) for SLE patients with neutralising (IC50>100) and non-neutralising (IC50<100) anti-IFNα-auto-Abs, and anti-IFNα-auto-Ab negative patients. \**P*<0.05 by unpaired Student’s *t*-test with Welch’s correction. **b**, Principal component analysis (PCA) of anti-IFNα-auto-Ab titres, neutralising capacity, BILAG GS, circulating IFNα concentration and disease duration in 54 SLE patients. Point colours indicate flares (BILAG score A/B) in different organ systems; central nervous system (CNS), cardiovascular (CVS), general (Gen), mucocutaneous, musculoskeletal (Mu/Skl), renal and none. **c, d**, Longitudinal analysis of (**c)** 2 SLE patients with neutralising anti-IFNα-auto-Abs and **(d)** 2 SLE patients with non-neutralising anti-IFNα-auto-Abs, showing titres of auto-Abs, circulating IFNα concentrations and BILAG GS. **e**, pSTAT stimulation index of monocytes treated with SLE patient sera containing IFNα and low titres of anti-IFNα-auto-Abs with and without a commercially available monoclonal anti-IFNα antibody, with sera from healthy controls, or with recombinant human (rh) IFNα2. \*\**P*=0.0035 by unpaired two-tailed *t*-test.

Principal component analysis (PCA) performed on 54 patients with detectable anti-IFNα-auto-Abs confirmed that the neutralizing capacity correlates with antibody titres, and that patients with high titres of neutralizing Ab clustered separately from those with active disease and high levels of serum IFNα (Fig. 2b). No specific organ involvement was identified amongst the different clusters of active patients. No significant differences in disease duration are found.

The results so far suggest that the presence of neutralizing anti-IFNα-auto-Abs is associated with a better clinical outcome. To gain more understanding into the kinetics of antibody production, putative reduction in IFNα levels and the effect on disease progression, we measured the levels of IFNα and anti-IFNα-auto-Ab titres in a longitudinal cohort of SLE patients. In 4 out of 5 patients (depicted here), high titres of neutralizing anti-IFNα-auto-Abs were stable over time, neutralized all the IFNα subtypes (with some variation with IFNα1) and reduced pan-IFNα protein levels in circulation down to undetectable levels (Fig. 2c, Supplementary Figure 2a-b). The prolonged presence of neutralizing anti-IFNα-auto-Abs together with a consistently low concentration of IFNα, mirrored a persistent inactive clinical score (Fig. 2c, Supplementary Figure 2a). Patients with non-neutralizing anti-IFNα-auto-Abs in circulation were characterized by high levels of IFNα and were generally associated with a more severe disease activity (Fig. 2d). These results suggest that these non-neutralising auto-Abs may stabilize circulating IFNα levels as previously suggested for other cytokines^28–30^. The association of non-neutralizing auto-Abs with high IFNα concentration is intriguing. It has been previously suggested that in certain cases, including more recently in COVID-19 patients^31^, circulating auto-antibodies can increase the half-life of the molecule they bind, possibly through the uptake and release of immune complexes by the neonatal Fc receptor on endothelial cells^32,33^.

It is plausible that IFNα bound to auto-Abs can still be detected by the Simoa assay (Bondet et al, submitted). Depending on the epitope bound, IFNα in immune complexes can be neutralized or remain bioactive. To assess whether IFNα in patients containing non-neutralizing auto-Abs is functionally active, we tested SLE patient sera on isolated monocytes using a pSTAT1 induction assay. pSTAT1 was upregulated by SLE sera containing both IFNα and low-titre anti-IFNα auto-Abs, in comparison to healthy control sera or patients without measurable IFNα or anti-IFNα auto-Abs. Excessive pSTAT1 upregulation was partially abrogated by preincubation with a monoclonal anti-IFNα antibody (26B9) that neutralizes all subtypes of IFNα (Fig. 2e). Taken together, our results suggest that anti-IFNα-auto-Abs influence disease outcome of SLE depending on the titres of antibodies and their neutralization capacity.

SLE patients are known to present a variety of B cell abnormalities, including increased frequencies of double-negative B cells and plasmablasts^1–3^. Previous work by us and others have demonstrated that the level of exposure to IFNα is crucial in determining immature B cell fate^6,7,34^. Whereas exposure of immature B cells to low-moderate concentrations of IFNα simultaneously expand both Bregs and plasmablasts, high concentrations of IFNα (observed in SLE patients) biased B cell differentiation towards pro-inflammatory plasmablasts and plasma cells^7^. In order to evaluate whether the presence of anti-IFNα-auto-Abs is associated to a normalization of the B cell frequencies and responses, we quantified *ex vivo* B cell subset frequencies in patients with and without anti-IFNα-auto-Abs followed by stratification according to their neutralizing capacity.

We observe a significant reduction in unswitched IgM memory (USM, IgD^+^CD27^+^) and a significant increase in DN (IgD^-^CD27^-^) Bcells and plasmablasts (CD19^low^CD27^+^IgD^-^CD38^hi^) in anti-IFNα-auto-Ab negative patients compared to healthy individuals (Fig. 3a-c). Our data also show a significant increase of immature B cells (CD24^hi^CD38^hi^) and a trend towards increase in frequencies of plasmablasts in the anti-IFNα-auto-Ab negative group compared to the positive one. Further stratification of anti-IFNα-auto-Ab positive SLE patients into neutralizing (IC50>100) and non-neutralising (IC50<100) groups revealed a significant increase of double negative memory B cells (DN, IgD^-^CD27^-^) in patients with non-neutralizing anti-IFNα-auto-Abs (Fig. 3d).

**Fig. 3:**
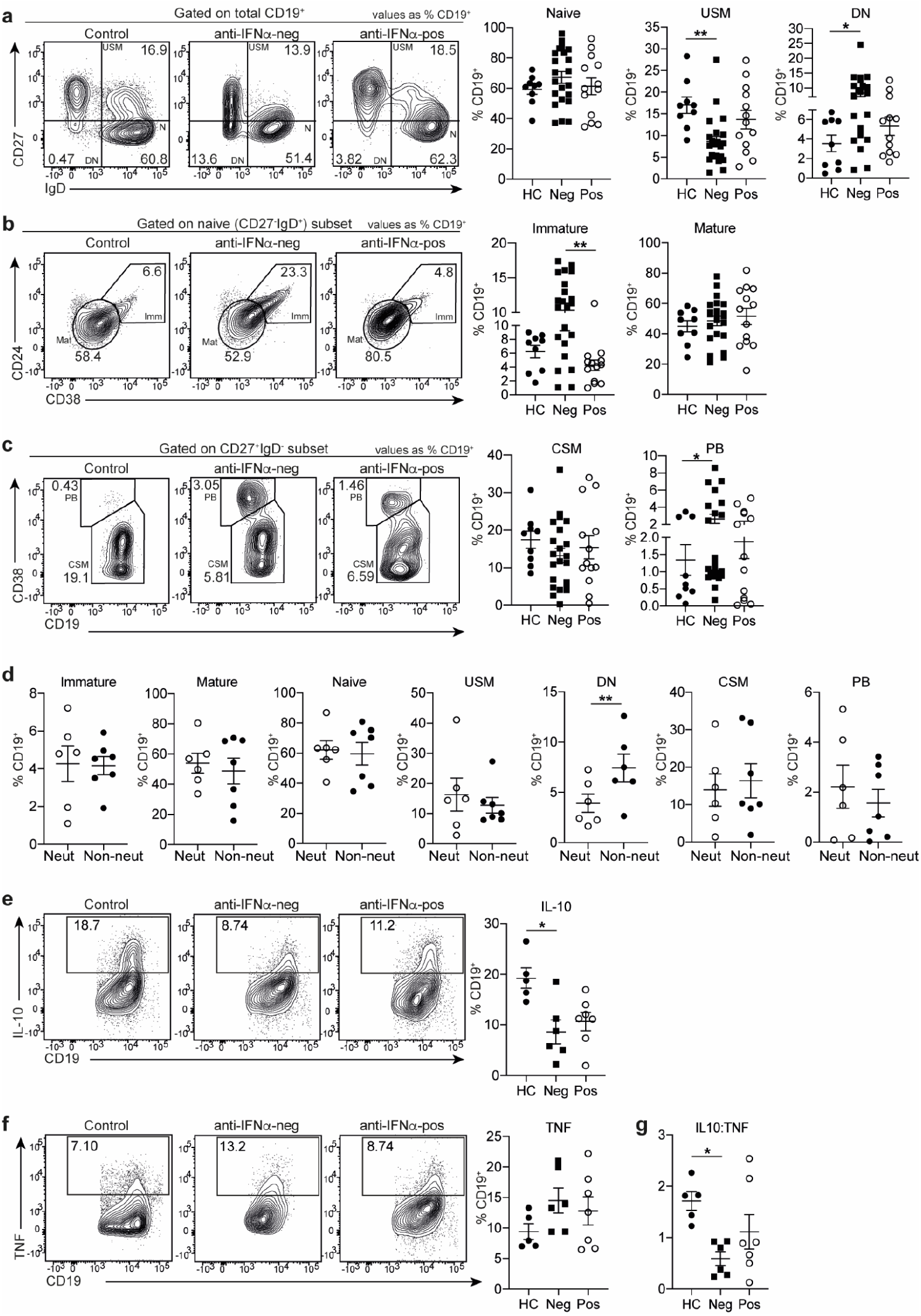
SLE patients with neutralizing anti-IFNα autoantibodies display normalized frequencies of peripheral blood B cell subsets. **a-c**, Representative contour plots and graphs show for anti-IFNα-auto-Ab positive (n=13), anti-IFNα-auto-Ab negative (n=22) SLE patients and healthy controls (n=9), *ex vivo* frequencies of **(a)** IgM^+^ unswitched memory (USM, CD27^+^ IgD^+^), double negative (DN, CD27^-^IgD^-^) and naïve (N, CD27^-^IgD^+^) B cells, **(b)**, immature (Imm, CD24^hi^CD38^hi^) and mature (Mat, CD24^int^CD38^int^) B cells gated within the naïve (CD27^-^IgD^+^) subset, and **(c)** class-switched memory B cells (CSM) and plasmablasts (PB) gated within the CD27^+^IgD^-^ subset. All values are given as % of total CD19^+^ population (gating strategy in Supplementary Fig. 3a). \**P*<0.05, \*\**P*<0.01 by non-parametric Kruskal-Wallis test with Dunn’s multiple comparison. **d**, Graphs show frequencies of B cell subsets as described in Fig. 3a-c in SLE patients with neutralizing (IC50>100) or non-neutralizing (IC50<100) anti-IFNα-auto-Abs. \*\**P*<0.01 by unpaired Student’s *t* test with Welch’s correction. **e-g**, Representative contour plots and graphs show frequencies of **(e)** IL-10^+^ and **(f)** TNF^+^B cells within the total CD19^+^ population and **(g)** ratios of IL-10^+^ to TNF^+^B cell frequencies following 72h CpGC stimulation of PBMCs isolated from anti-IFNα-auto-Ab positive (n=7), anti-IFNα-auto-Ab negative (n=6) SLE patients and healthy controls (n=5). \**P*<0.05 by non-parametric Kruskal-Wallis test with Dunn’s multiple comparison. Error bars are shown as mean±SEM. Data in e and f are representative of at least 3 independent experiments.

Bregs (hereafter defined as IL-10^+^B cells) are numerically and functionally impaired in SLE patients^35^, partially as a consequence of excessive IFNα production by pDCs skewing B cells towards plasmablast differentiation^7^. To establish whether SLE patients with anti-IFNα-auto-Abs regain IL-10^+^B cells differentiation capacity we stimulated PBMCs from SLE patients and healthy controls with CpGC for 72h to induce both pDC IFNα production and IL-10^+^B cells differentiation. There is a significant decrease in IL-10^+^B cells in anti-IFNα-auto-Ab negative patients but not in anti-IFNα-auto-Ab positive patients compared to healthy individuals (Fig. 3e). No differences in frequencies of TNFα+ B cells were observed in anti-IFNα-auto-Ab negative or anti-IFNα-auto-Ab positive patients compared to healthy controls (Fig. 3f). Therefore, the net outcome results in a significantly decreased ratio of IL-10^+^B cells to TNFα+B cells in anti-IFNα-auto-Ab negative but not positive patients compared to healthy controls (Fig. 3g). Our findings show that there are no differences in the frequencies of T follicular helper cell (TFH) subsets (circulating (cTFH) or activated (aTFH) between patients and healthy controls (Supplementary Figure 3b-c). This supports a direct role of anti-IFNα-auto-Abs in normalising B cell subset frequencies rather than indirectly via modifications to the TFH compartment.

In response to viral infections, plasmacytoid dendritic cells (pDCs) rapidly produce IFNα that drives B cell maturation into plasma cells producing antibody against viral antigens^6^. In view of the recent findings showing the detrimental effect of neutralizing anti-IFNα-auto-Abs in patients with COVID-19, it is important to understand the impact of neutralizing anti-IFNα-auto-Abs on “nascent” IFNα produced by challenged pDCs, and how this affects healthy B cell differentiation. PBMCs from healthy donors were stimulated with CpGC and cultured respectively with serum from SLE patients with no antibodies or with neutralizing anti-IFNα-auto-Abs; healthy allogeneic serum was used as a control (as depicted in Fig. 4a). Sera from patients containing neutralizing anti-IFNα-auto-Abs significantly downregulated ISG expression in cultured healthy PBMCs confirming their ability to inhibit IFNα-down-stream signalling (Fig. 4b, Supplementary Figure 3d). Although sera from both groups of SLE patients decreased to a certain extent the frequency of immature B cells and plasmablasts and increased memory and mature B cells, the sera from anti-IFNα-auto-Ab positive patients profoundly reduced healthy immature B cell and plasma cell differentiation confirming the *ex vivo* neutralizing capacity of anti-IFNα-auto-Abs (Fig. 4c).

**Fig. 4:**
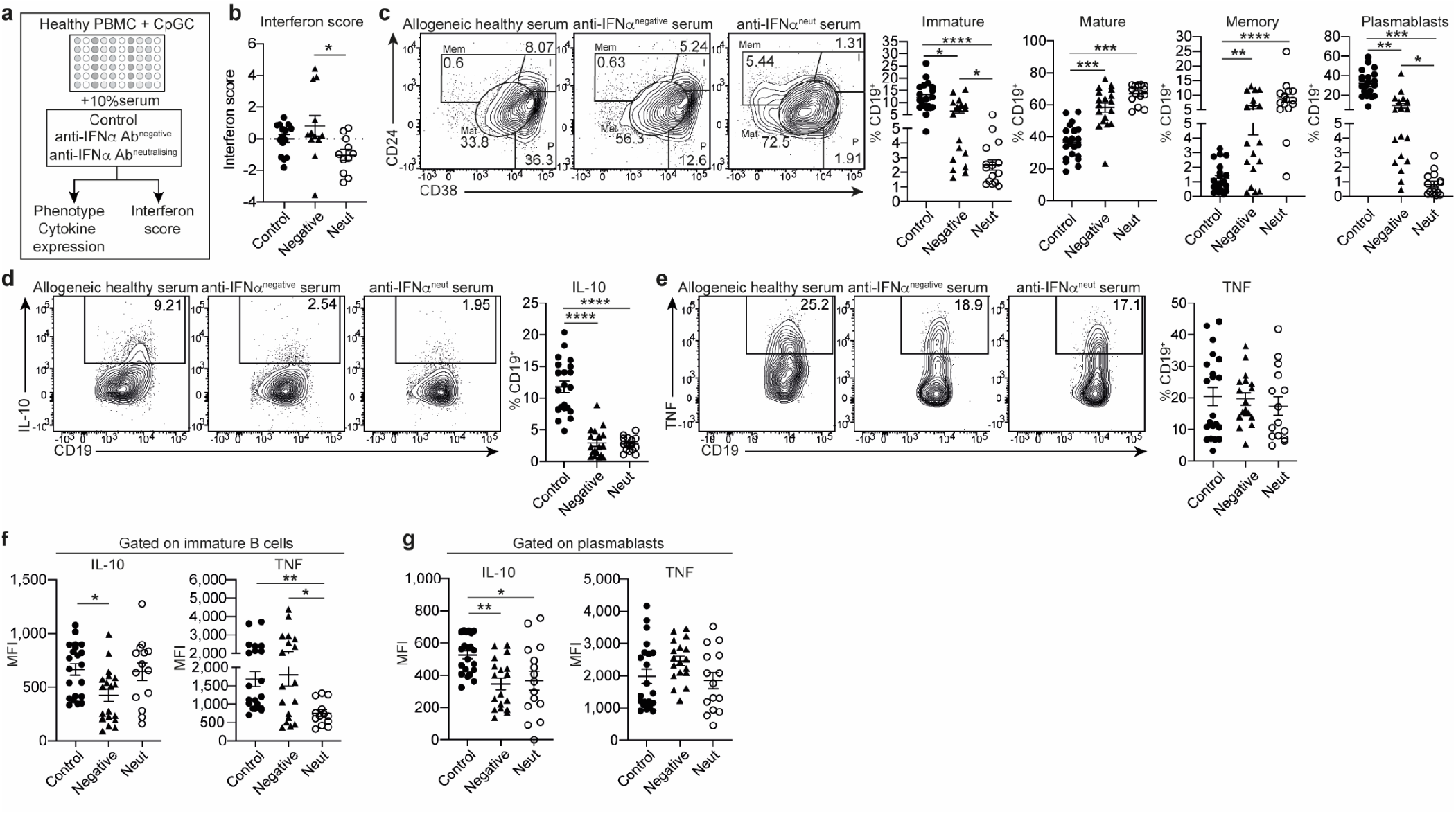
SLE serum with neutralising anti-IFNα-autoantibodies inhibit IFNα-induced immature B cell and plasmablast differentiation. **a**, Schematic of serum transfer experimental design; healthy PBMCs were cultured with CpGC and 10% serum from SLE patients with neutralizing (IC50>100) anti-IFNα-auto-Abs, from anti-IFNα-auto-Ab negative SLE patients and healthy controls for 72 hrs. **b**, Interferon score of PBMCs following culture in SLE or healthy control sera. Data represented are a cumulative score of the expression of ISGs MX1, MCL1, IRF9 and STAT1 measured by RT-qPCR and relative to the GAPDH **c-e**, Representative contour plots and graphs showing frequencies of **c**, immature (I, CD24^hi^CD38^hi^), mature (Mat, CD24^int^CD38^int^), total memory (Mem, CD24^+^CD38^lo^) B cells and plasmablasts (P, CD24^lo/-^ CD38^hi^), **d**, IL-10^+^ and **e**, TNF^+^B cells. Values are given as % total of the CD19^+^ population. **f, g**, Mean fluorescence intensities (MFI) of IL-10 and TNF expression within **(f)** immature and (**g)** plasmablast subsets. *\*P*<0.05, \*\**P*<0.01, \*\*\**P*<0.001, \*\*\*\**P*<0.0001 by non-parametric Kruskal-Wallis test with Dunn’s multiple comparison. Error bars are shown as mean±SEM. Data are representative of 2 independent experiments.sssw

The frequencies of IL-10^+^B cells is significantly decreased after culture with both groups of SLE sera compared to control sera (Fig. 4d). There is no difference observed in the frequency of TNFα+B cells (Fig. 4e).

Amongst different B cell subsets that produce IL-10, immature B cells and plasma cells are the two subsets that produce the most of IL-10. It is interesting, to note that the remaining immature B cells present in the culture stimulated with the serum from the anti-IFNα-auto-Ab positive patients, produced similar levels of IL-10 as B cells stimulated with control sera (Fig. 4f). Expression of IL-10 by the plasmablast subset was significantly reduced by both anti-IFNα-auto-Ab positive and negative sera compared to control (Fig. 4g). The overall reduced frequencies of IL-10^+^B cells in PBMCs treated with sera containing neutralising anti-IFNα-auto-Abs may be, on one hand, a reflection of the contraction of the immature B cell compartment, since IL-10^+^B cells have been shown to be enriched within this subset following IFNα stimulation^35^. However, it should be also acknowledged that sera from anti-IFNα auto-Ab negative patients significantly inhibited IL-10^+^B cell expansion, suggesting that other unknown factors, present in the sera of SLE patients, contribute to the overall observed inhibition of IL-10^+^B cells.

In summary, we report that a subset of SLE patients harbour neutralizing anti-IFNα-auto-Abs that can modulate B cell responses and are associated to a better disease outcome. This is in contrast with patients with non-neutralizing low titres of anti-IFNα-auto-Abs, which appear to stabilize IFNα in the blood and expand circulating frequencies of DN memory B cells and plasmablasts B cells. Future studies investigating why only a subset of SLE patients develop high titres of neutralizing anti-IFNα-auto-Abs while others develop lower, non-neutralizing titres are warranted. Our findings are particularly relevant in the current COVID-19 pandemic, where anti-IFN-I-auto-Abs and impaired IFN signalling have been associated with higher susceptibility for serious illness^23^. Thus, SLE patients with broadly neutralizing anti-IFNα-auto-Abs could potentially be at high risk of developing life-threatening COVID-19 and for these reasons their vaccination should be prioritized, and the antibody responses carefully monitored.

## Data Availability

No data sets were generated or analysed during the current study.

## Materials and methods

### Patients and controls

Blood samples for PBMC and serum isolation were collected from SLE patients attending the University College London Hospital (UCLH) rheumatology clinic, and from healthy volunteers following informed consent. Ethical approval was obtained from the UCLH Health Service ethical committee, under REC reference no. 14/SC/1200. Sample storage complied with requirements of the Data Protection Act 1998.

### PBMC and serum isolation

A total of 50ml whole peripheral blood was collected from an individual patient or healthy donor for PBMC isolation using Ficoll-based density gradient centrifugation. A total of 10ml whole peripheral blood was collected into serum-separator (SST) tubes, centrifuged for 10 minutes at 1200g at RT and serum decanted.

### Luciferase Immunoprecipitation system (LIPS) assay

LIPS was performed as previously described (Meyer *et al*., 2016). Briefly, different IFNα subtype and cytokine sequences were cloned into modified pPK-CMV-F4 fusion vector (PromoCell GmbH, Germany) where Firefly luciferase was substituted in the plasmid for *NanoLuc* luciferase (Promega, USA). Cloned constructs were transfected into HEK293 cells, and after 48h tissue culture media containing fusion proteins were collected and stored at −20°C. IgG from the serum samples was captured onto Protein G Agarose beads (Exalpha Biologicals, USA) at room temperature for 1h in 96-well microfilter plate (Merck Millipore, Germany). Antigens were added to microfilter plate at 1×10^6^ luminescence units (LU) per well and incubated at room temperature for 1h. After washing the plate with vacuum system, Nano-Glo® Luciferase Assay Reagent was added (Promega, USA). Luminescence intensity was measured by VICTOR X Multilabel Plate Reader (PerkinElmer Life Sciences, USA). The results were expressed as arbitrary units (AU) representing the percent of signal intensity from a positive control sample. Positve negative discrimination level was calculated as mean plus 3 SD from 1% trimmed values of healthy controls.

For the detection of IgG subclass-specificity, serum samples were incubated with fusion protein solutions (10^6^ LU per well) overnight at +4°C. Next day, agarose beads bound with streptavidin (Novagen, USA) were incubated with biotin-conjugated human subclass-specific antibodies (anti-IgG1, anti-IgG2, anti-IgG4 from BD Pharmingen, USA; anti-IgG3 from Sigma-Aldrich, USA) in microfilter plates for 1 h at room temperature. Overnight incubated serum samples with fusion protein solutions were added to microfilter plate and incubated at room temperature for 2h. Microfilter plates were washed and luminescence intensity measured as above. The results were expressed as luminescence units (LU).

### Neutralization assay

Type I interferon neutralizing capacity was measured by using a reporter cell line HEK-Blue IFN-α/β (InvivoGen, USA) as previously described (Meyer *et al*., 2016). The cells were grown in DMEM (Lonza, Switzerland) with heat-inactivated 10% FBS, 30 g/ml Blasticidin (InvivoGen, USA) and 100 g/ml Zeocin (InvivoGen, USA). IFN-α2 was used at concentration 25 U/ml (Miltenyi Biotech, Germany). Serial dilutions were made to find the optimal dilution for other IFN-α subtypes (IFN-α1, IFN-α4, IFN-α5, IFN-α6, IFN-α7, IFN-α8, IFN-α10, IFN-α14, IFN-α16, IFN-α17, IFN-α21 from PBL Assay Science, USA). The dilution that induced approximately the same alkaline phosphatase (AP) concentration as IFN-α2 25 U/ml was selected for further neutralization assays.Three-fold serially diluted serum samples were co-incubated with interferons for 2h at 37°C, 5% CO2. 10^5^ IFN-α-HEK-Blue cells were added to microtiter plate wells and incubated 20-24hours at 37°C, 5% CO2. QUANTI-Blue (InvivoGen, USA) colorimetric enzyme assay was used to determine AP activity in overnight supernatants. Optical density (OD) was measured at 620 nm with Multiscan MCC/340 ELISA reader (Labsystems, USA). Neutralization activity was expressed as IC50, which was calculated from the dose-response curves and represents the serum dilution at which the IFN bioactivity was reduced to half of its maximum.

### pSTAT1 induction assay

Monocytes were stimulated with IFNα (dilution series between 12.5 U/ml to 400 U/ml) (Miltenyi Biotec), with two SLE patient serums with IFNα concentration 11204 fg/ml and 2424 fg/ml and low levels of anti-IFNα auto-Abs, and 6 control serum samples. Monocytes were stimulated for 20 minutes, fixed with BD Cytofix Fixing buffer (BD Biosciences), permeabilized with Perm Buffer III (BD Biosciences) and stained with p-STAT1 antibody (BD Phosflow Alexa Fluor 647 mouse anti-STAT1, pY701, clone 4a, BD Biosciences) according to manufacturer’s protocol. After staining, cells were analyzed using a Digital LSR Fortessa flow cytometer (BD Biosciences) and FCS Express 5 Flow (*De Novo* Software).

### IFNα concentration measurement

Simoa digital ELISA was performed to measure IFNα concentration in patient serum as previously described^25^. Two IFNα specific antibodies (cloned from APECED patients) described previously (Meyer et al., 2016)] were used. The 8H1 antibody clone was used as a capture antibody after coating on paramagnetic beads (0.3 mg/ml), and the 12H5 was biotinylated (biotin/antibody ratio = 30/1) and used as the detector. The limit of detection was was 0.7 fg/ml. The results are expressed in fg/ml.

### Primary cell cultures

PBMCs were cultured in 96-well plates at a density of 5×10^6^ cells/ml in RPMI 1640 (Sigma-Aldrich) supplemented with 10% FCS and 100 IU/mg penicillin/streptomycin (Sigma-Aldrich). PBMCs were stimulated with 1mM CpGC ODN 2395 (InvivoGen), then incubated for 72hrs at 37°C and 5% CO2. For serum transfer experiments, healthy PBMCs were stimulated with 1mM CpGC ODN 2395 and cultured in RPMI 1640 supplemented with 100 IU/mg penicillin/streptomycin and 10% serum from healthy donors and SLE patients with high, low or negative titres of anti-IFNα auto-Abs, with serum IFNα < 2 fg/ml.

### Flow cytometry staining and analysis

PBMCs were stained at a maximal concentration of 5×10^6^ cells/ml in staining buffer in the dark (PBS, 2% FCS 1mM EDTA) or as indicated below. For exclusion of dead cells from analysis, cells were incubated for 20 min with 1:500 Live/Dead Fixable Blue Dead Cell Stain Kit (ThermoFisher) at room temperature. Cell surface markers were stained with the following directly conjugated antibodies from BioLegend: CD19 PE/Cy7 (HIB19), IgM BV510 (MHM-88), IgD BV605 (IA6-2), CD27 PE/Dazzle 594 (M-T271). CD24 APCeFluor780 (SN3 A5-2H10) and CD38 PerCPeFluor710 (HB7) were purchased from eBioscience. NK/TFH staining was performed using the following directly conjugated antibodies; CD3 AlexaFluor488 (BioLegend, UCHT1), CD4 PE/Dazzle594 (BioLegend, A161A1), CD8 BUV395 (BD, RPA-T8), CD56 APC-Cy7 (BioLegend, NKH1), CXCR5 BV421 (BioLegend, J252D4), ICOS PE/Cy7 (BioLegend, 2D3), PD-1 BUV737 (BD, EH12.1), CCR7 BV785 (BioLegend,G043H7). CD159c-Biotin (Miltenyi, REA205) was conjugated to Streptavidin-BV510 (BioLegend). For multi-colour flow cytometric surface marker analysis cells were stained for 30 min at 4°C. Cells were incubated for 10 min at 4°C in fixation buffer containing formaldehyde (eBioscience).

For intracellular cytokine staining of cultured cells, cells were stimulated with PMA (50 ng/ml), ionomycin (250 ng/ml) and brefeldin A (5mg/ml) for the final 5 hours of culture. Surface markers and dead cells were stained as previously described. Following fixation cells were permeabilised (eBioscience) and incubated with IL-10 APC (BD, JES5-16E3), IL-6 PE (Biolegend, MQ2-13A5) and TNF eFluor450 (eBioscience, mAb11), or PLZF APC (eBioscience, 9E12) for 40 mins. Cells were acquired using a Digital LSR II flow cytometer (Becton Dickinson). Flow cytometric data were analysed with FlowJo software v.10.4.1 (TreeStar).

### RT-PCR

Total RNA was extracted from total PBMCs using the Arcturus PicoPure kit (ThermoFisher) and RNase-Free DNase Set (Qiagen) as per manufacturer’s instructions. RNA was reverse transcribed to cDNA using the iScript cDNA synthesis kit (Bio-Rad) and gene expression measured by quantitative RT-PCR. PCR primers used are as follows; MCL1, IRF9, MX1, STAT1 (QIAGEN), and GAPDH (forward, 5’ CGCTCTCTGCTCCTCCTGTT, reverse 5’ GCAAATGAGCCCCAGCCTTCTC). For interferon scores, ISG relative expression values were summed and score calculated as the number of standard deviations (of summed values from healthy donors; SD(HD)) above the mean of summed healthy donor values; MEAN(HD). Cut-off values were calculated as the MEAN(HD) + 2SD(HD).

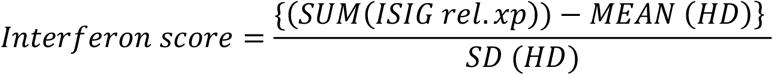

## Statistical analysis

Statistical analysis was performed with Prism Software (GraphPad), using unpaired t tests or non-parametric analysis using the Kruskal-Wallis test with Dunn’s multiple comparison test for multiple comparisons. Correlations were assessed with Pearson’s correlation coefficient. A p value of < 0.05 was considered as significant. ns: not significant, *p < 0.05, **p < 0.01, ***p < 0.001, ****p<0.0001.

## Data availability statements and data citations

No data sets were generated or analysed during the current study.

## Acknowledgements

This work is funded by Versus Arthritis UK program grant (21140) and Research Award (21786) to C.M., by the European Regional Development Fund (Project No 2014-2020.4.01.15-0012 and the Centre of Excellence in Genomics (EXCEGEN) framework) (LH, PP, KK), the Estonian Research Council grants PRG1117 (KK) and PRG377 (PP). H.F.B. is funded by a UCB BIOPHARMA SPRL/BBSRC PhD Studentship (BB/P504725/1). We thank Immunoqure for provision of mAbs for the pan-IFNα assay under an MTA to D.D. D.D. acknowledges support from the ANR (CE17001002). We thank Drs. Diego Catalan and Christopher Piper for critically reviewing the manuscript.

## Supplementary Figures

**Supplementary Fig. 1.**
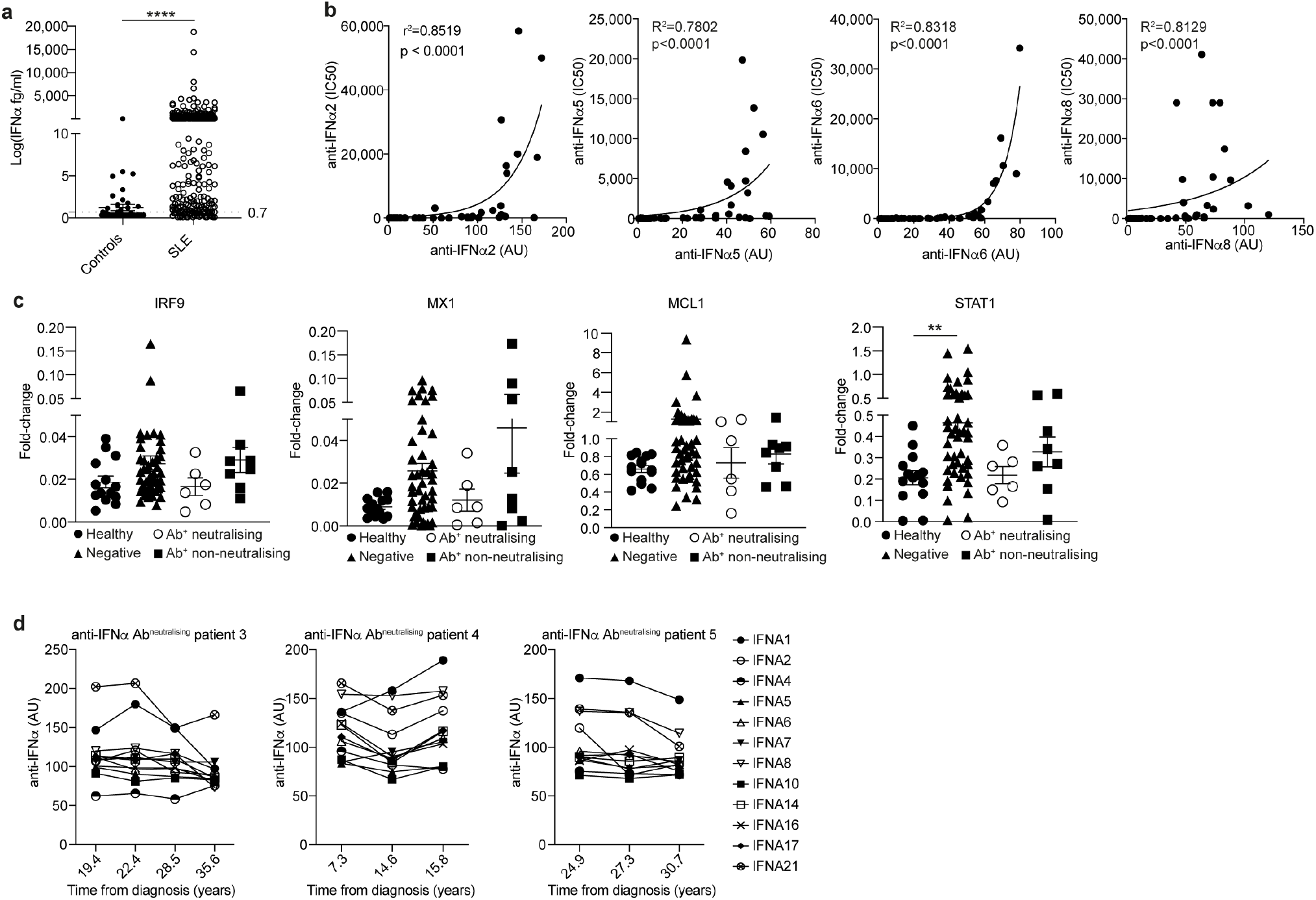
**a,** Serum IFNa levels (fg/ml) for 54 healthy controls and 470 SLE patients. \*\*\*\**P*<0.0001 by unpaired Student’s *t* test with Welch’s correction. **b**, Correlations between anti-IFNα-auto-Ab titres and neutralizing capacity (IC50) for auto-Ab subtypes IFNα2, IFNα5, IFNα6 and IFNα8. \*\*\*\**P*<0.0001 by two-tailed nonparametric Spearman correlation. **c**, Titres of anti-IFNα-auto-Abs against different IFNα subtypes longitudinally for 3 SLE patients with neutralizing anti-IFNα-auto-Abs. **d**, Fold-change expression of individual ISGs IRF9, MX1, MCL1 and STAT1 for PBMCs isolated from SLE patients with neutralizing or non-neutralizing anti-IFNα-auto-Abs, anti-IFNα-auto-Ab^-^ SLE patients and healthy controls, as measured by RT-qPCR. \*\**P*=0.0031 by non-parametric Kruskal-Wallis test with Dunn’s multiple comparison. Error bars are shown as mean±SEM.

**Supplementary Fig. 2.**
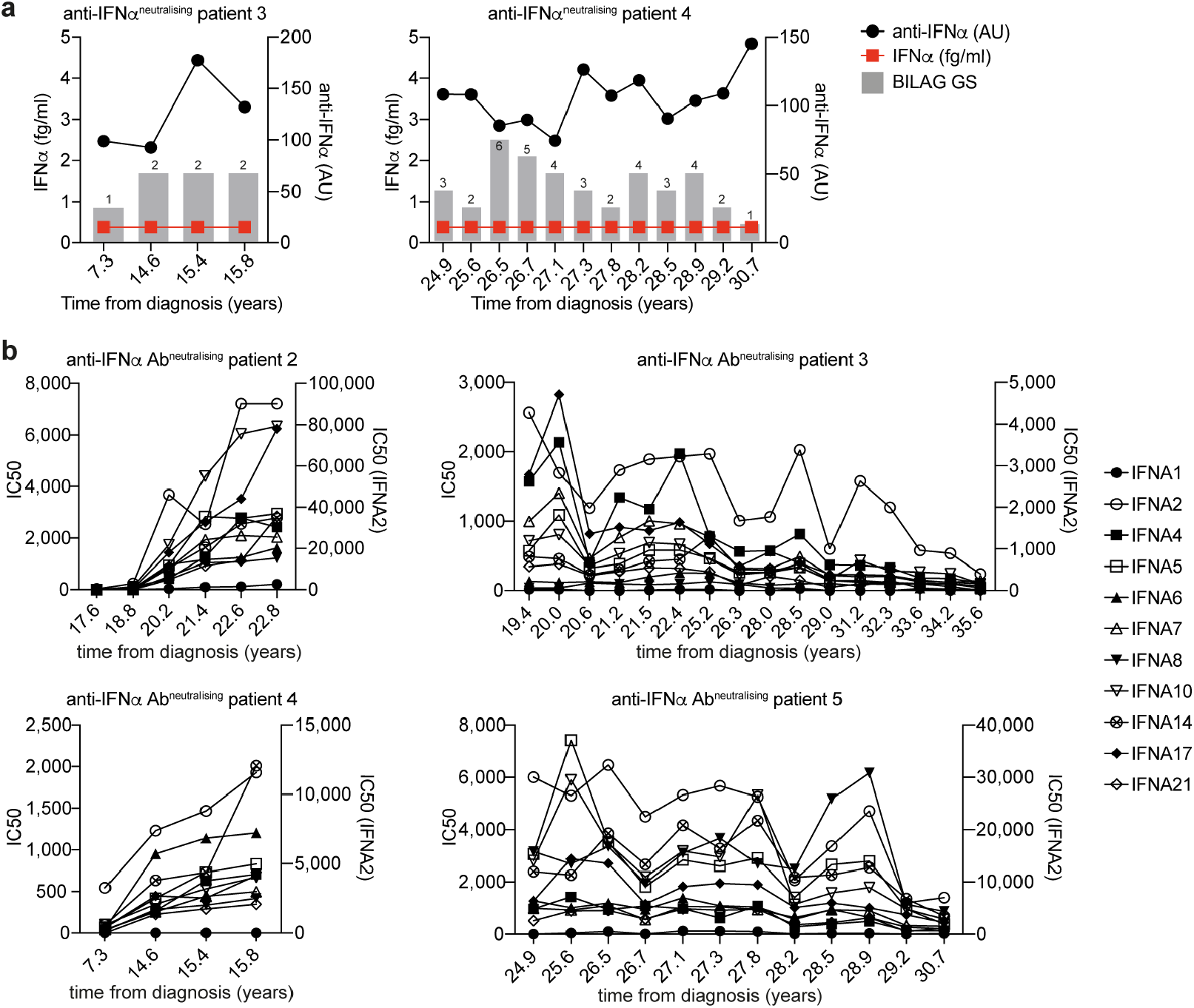
**a,** Longitudinal analysis of 2 SLE patients with neutralising anti-IFNα-auto-Abs showing titres of auto-Abs, circulating IFNα concentrations and BILAG GS. **b**, Longitudinal analysis of 4 SLE patients showing neutralising capacity of anti-IFNα-auto-Abs against individual IFNα subtypes IFNα1/4/5/6/7/8/10/14/17/21 (left y axis) and IFNα2 (right y axis).

**Supplementary Fig. 3.**
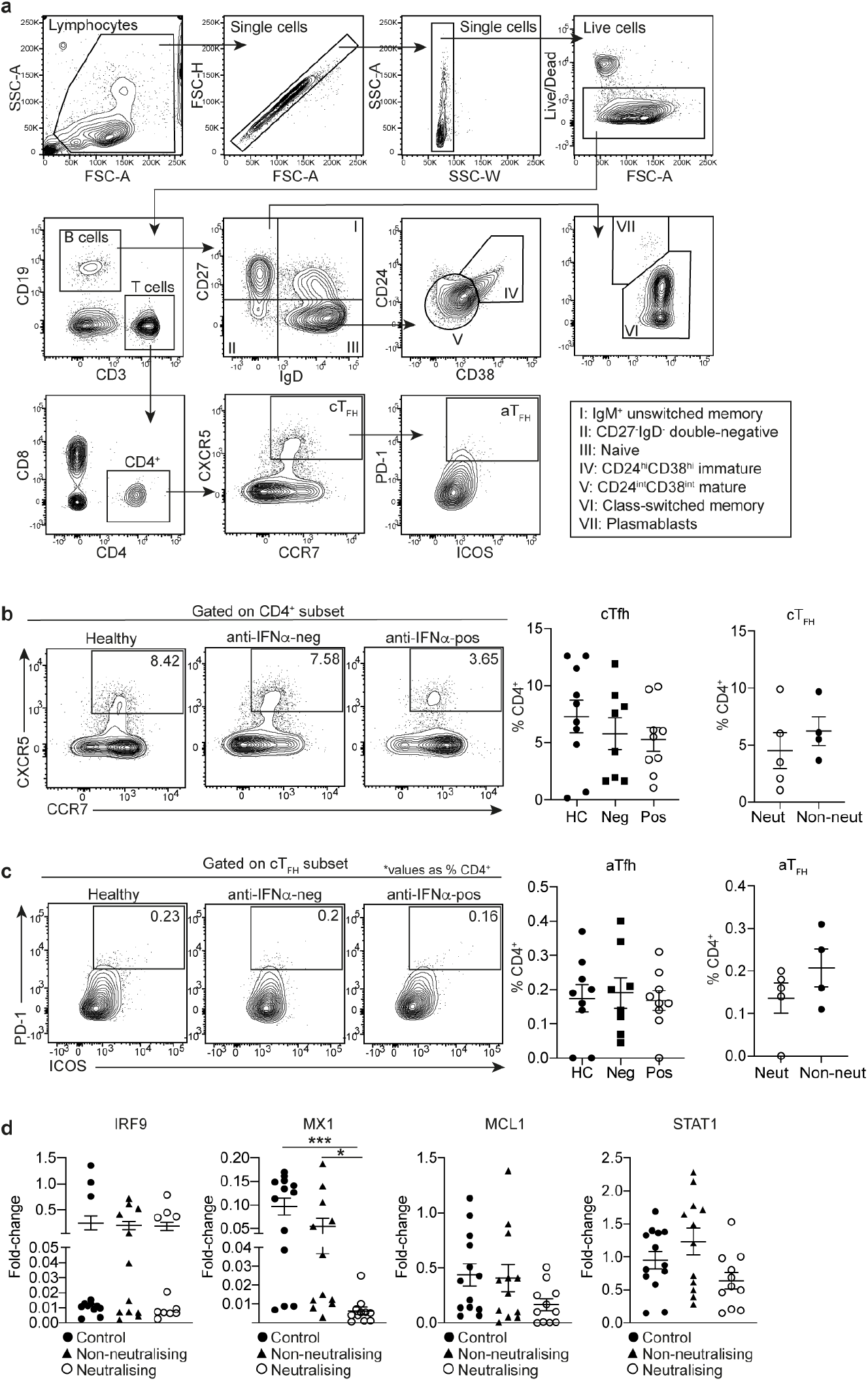
**a,** Gating strategy for *ex vivo* measurement of B cell subset and TFH subset frequencies by flow cytometry. **b-c**, Representative contour plots and graphs show *ex vivo* frequencies of **(b)** classical TFH (cTFH) and **(c)** activated TFH (aTFH) T cells in anti-IFNα auto-Ab positive and anti-IFNα-auto-Ab negative SLE patients and healthy controls. Values are given as % total CD4^+^ population. **d**, Fold-change expression of individual ISGs IRF9, MX1, MCL1 and STAT1 for PBMCs following 72h culture with CpGC and 10% serum from SLE patients with neutralizing anti-IFNα-auto-Abs, anti-IFNα-auto-Ab negative and healthy controls, as measured by RT-qPCR. \**P*<0.05, \*\*\**P*<0.001 by nonparametric Kruskal-Wallis test with Dunn’s multiple comparison. Error bars are shown as mean±SEM.

